# Ambient nitrogen dioxide in 47,187 neighborhoods across 326 cities in eight Latin American countries: population exposures and associations with urban features

**DOI:** 10.1101/2023.05.02.23289390

**Authors:** Josiah L. Kephart, Nelson Gouveia, Daniel A. Rodriguez, Katy Indvik, Tania Alfaro, José Luis Texcalac, J. Jaime Miranda, Usama Bilal, Ana V. Diez Roux

## Abstract

**Background:** Health research on ambient nitrogen dioxide (NO_2_) is sparse in Latin America, despite the high prevalence of NO_2_-associated respiratory diseases in the region. This study describes within-city distributions of ambient NO_2_ concentrations at high spatial resolution and urban characteristics associated with neighborhood ambient NO_2_ in 326 Latin American cities.

**Methods:** We aggregated estimates of annual surface NO_2_ at 1 km^2^ spatial resolution for 2019, population counts, and urban characteristics compiled by the SALURBAL project to the neighborhood level (i.e., census tracts). We described the percent of the urban population living with ambient NO_2_ levels exceeding WHO Air Quality Guidelines. We used multilevel models to describe associations of neighborhood ambient NO_2_ concentrations with population and urban characteristics at the neighborhood and city levels.

**Findings:** We examined 47,187 neighborhoods in 326 cities from eight Latin American countries. Of the ≈236 million urban residents observed, 85% lived in neighborhoods with ambient annual NO_2_ above WHO guidelines. In adjusted models, higher neighborhood-level educational attainment, closer proximity to the city center, and lower neighborhood-level greenness were associated with higher ambient NO_2_. At the city level, higher vehicle congestion, population size, and population density were associated with higher ambient NO_2_.

**Interpretation:** Almost nine out of every 10 residents of Latin American cities live with ambient NO_2_ concentrations above WHO guidelines. Increasing neighborhood greenness and reducing reliance on fossil fuel-powered vehicles warrant further attention as potential actionable urban environmental interventions to reduce population exposure to ambient NO_2_.

**Funding:** Wellcome Trust, National Institutes of Health, Cotswold Foundation

## 1. Introduction

Ambient nitrogen dioxide (NO_2_) is a ubiquitous urban air pollutant produced by fossil fuel combustion. NO_2_ is emitted by outdoor and indoor point sources, such as industrial processes and household cooking and heating,(1) and through mobile sources, such as exhaust from fossil fuel-powered vehicles(1). Under certain atmospheric conditions, NO_2_ can rapidly transform to other chemical structures and is a key ingredient in the formation of ground-level ozone(1). Because of NO_2_’s tendency to rapidly transform over time and space, ambient NO_2_ concentrations can have highly granular spatial variability within cities(2, 3, 4). This variability is often linked to spatially-varying social characteristics, resulting in within-city social disparities in NO_2_ exposures(5).

Historically, epidemiologic research on NO_2_ has faced the challenge of examining NO_2_ exposure within complex mixtures of co-occurring pollutants, such as exhaust from fossil fuel-powered vehicles, and many epidemiologic analyses have approached ambient NO_2_ as a proxy for traffic-related air pollutant mixtures(4). However rapidly growing evidence supports increased attention to the role of NO_2_ itself as an independent risk factor for health(6). Exposure to NO_2_ contributes to respiratory disease(1) and all-cause mortality(7, 8, 9, 10), among other health effects(1).

Children, the elderly, and individuals with respiratory disease are particularly susceptible to the health effects of NO_2_ exposure(1). In 2021, the WHO lowered the Air Quality Guideline for annual NO_2_ by 75% (from 40 ug/m3 to 10 ug/m3), citing growing evidence of the impacts of NO_2_ on health(11). The updated WHO air quality guidelines and, specifically, the substantial reductions in guidelines for NO_2_ warrants renewed attention towards who is exposed to harmful ambient NO_2_ concentrations and how ambient NO_2_ can be reduced in urban settings that remain highly dependent on fossil fuels for transit and industry.

To date, limited research has examined population exposures to ambient NO_2_ and urban factors associated with ambient NO_2_ in Latin America. Even fewer studies have examined within-city differences in NO_2_ concentrations in the region. However, Latin America has both high urbanization (80% of the population lives in urban areas)(12) and a high prevalence of NO_2_-associated respiratory diseases(13). A 2019 study of asthma incidence attributable to ambient NO_2_ found that Lima, Peru and Bogotá, Colombia were among the top three cities globally for asthma incidence attributable to NO_2_ exposure(13). A 2022 study of 968 urban areas in Latin America estimated that 16% of pediatric asthma cases in Latin American cities are attributable to ambient NO_2_ exposures(14). Despite the substantial health impacts of ambient NO_2_ in the region, ambient NO_2_ monitoring networks in the region are sparse(15). However, recent advances in satellite-derived global estimates of surface NO_2_ at fine spatial resolution(14) provide novel opportunities to examine social disparities and spatial variations in population exposures to NO_2_ within and between cities in this highly urbanized region.

To address these knowledge gaps on population exposures to ambient NO_2_ in Latin America and the relationship between NO_2_ and the urban environment, this study aims to describe population exposures to ambient NO_2_ and urban characteristics associated with differences in ambient NO_2_ exposure at the census tract level within 326 Latin American cities.

## 2. Methods

### 2.1. Study setting

This study was conducted as part of the *Salud Urbana en América Latina* (SALURBAL) project. The SALURBAL study protocol was approved by the Drexel University Institutional Review Board (ID no. 1612005035). This international scientific collaboration has compiled and harmonized data on social, environmental, and health characteristics for hundreds of cities in 11 Latin American countries(16). Cities in SALURBAL are composed of clusters of administrative units (i.e., municipalities) encompassing the visually apparent urban built-up area as identified using satellite imagery(17). Cities were defined as all urban agglomerations within the 11 countries that contained more than 100,000 residents as of 2010(17), facilitating examination of a diverse set of cities, from small cities to megacities. The SALURBAL project previously published an analysis of the variability and predictors ambient fine particulate matter (PM_2.5_) across the Latin American region (18), focusing on larger administrative units (i.e., municipalities).

In this analysis, we examine neighborhood-level ambient NO_2_ in 326 cities in Argentina, Brazil, Chile, Colombia, Costa Rica, Guatemala, Mexico, and Panama. Cities in El Salvador, Nicaragua, and Peru were excluded due to the lack of available data at the neighborhood level. Neighborhood administrative units varied in name and official definition by country. We used the country-specific, small-area administrative units most analogous to U.S. census tracts, henceforth referred to as “neighborhoods.” Detailed information on the administrative units and census used for each country is available in Supplementary Table S1. Across countries, these neighborhoods had a median population of 2,063 and a median area of 0.34 km^2^ (equivalent to a square with 0.58 km sides).

### 2.2. Neighborhood NO_2_ exposures

We used estimates published in 2022 of annual surface NO_2_ at 1 km^2^ spatial resolution(14). These estimates were based on a previous land use regression (LUR) model of mean surface NO_2_ from 2010-2012 at 100 m resolution(19). The LUR estimates were subsequently adjusted for bias using chemical transport models and scaled to an extended timeframe (annual means from 2005-2020) using satellite NO_2_ columns from the Ozone Monitoring Instrument v. 4.0 product(14, 20). For this analysis, we used NO_2_ estimates from the year 2019, the most recent year before the dramatic changes in ambient air pollution associated with the COVID-19 pandemic(21). To estimate neighborhood annual mean NO_2_, we averaged the values of all NO_2_ raster grid cells overlapping or contained within the neighborhood spatial boundary, area-weighting for the proportion of each grid cell contained within the neighborhood boundary. The resulting output was an estimate of annual surface NO_2_ in the year 2019 for each neighborhood in the study area.

### 2.3. Neighborhood and city characteristics

We used data on neighborhood characteristics and population compiled from national census bureaus and other sources by the SALURBAL project(17). We used the most recent available census for each country; information on the year of each census used is available in Supplementary Table 1. At both the neighborhood and city levels, the SALURBAL project previously estimated population density (population divided by built-up area), educational attainment (% of the population aged 25 years or older who completed primary education or above), intersection density (density of the set of nodes with more than one street emanating from them per km^2^ of built-up area), and area median greenness measured by the normalized difference vegetation index (NDVI). NDVI was calculated using MODIS satellite-based observations from the MODIS vegetation product, MOD13Q1.006 for 2015 at a 250 m spatial resolution(22). We computed the maximum NDVI value for 2019 at 250 m resolution to present the ‘greenest’ condition of each grid cell, then calculated the median across grid cells contained within each neighborhood. At the neighborhood level, we also calculated distance from the city center as the Euclidean distance (km) between the neighborhood centroid and city hall. At the city level, we also estimated city population, GDP per capita (computed as purchasing power parities in constant 2011 international USD of each city in 2015 using estimates from the first subnational administrative level, typically equivalent to departments or states(23)), and city-level traffic congestion (increase in road vehicle travel time due to congestion in the street network(24)).

### 2.4. Statistical analysis

We calculated summary statistics and boxplots of neighborhood NO_2_ concentrations, overall and stratified by country. Due to the small number of cities represented by each country, we pooled cities from Costa Rica (N=1 city), Guatemala (N=2), and Panama (N=3) into a single country grouping for Central America for all analyses. For comparability with the NO_2_ data source, we transformed the WHO annual guideline from μg/m^3^ to parts per billion (ppb; 10 μg/m^3^ ≈ 5.3 ppb) under standard assumptions (atmospheric pressure at sea level and 25°C temperature). We created summary statistics of the population living in neighborhoods with ambient NO_2_ levels above and below annual WHO Air Quality Guidelines (10 μg/m^3^), overall and by country. We summarized city-level mean NO_2_ for descriptive analysis by aggregating neighborhood NO_2_ estimates to the city-level using a population-weighted average.

To estimate between-country vs. between-city vs. within-city variation in neighborhood ambient NO_2_ exposures, we used a mixed effects one-way ANOVA with random intercepts for city and country.

We used multilevel univariable and multivariable models to describe associations between neighborhood-level ambient NO_2_ concentrations and population and urban characteristics at the neighborhood and city levels. All independent variables were operationalized as z-scores of the overall study distribution for each respective variable. We first conducted a univariable analysis of each independent variable and the dependent variable of neighborhood annual NO_2_. We then modeled all neighborhood- and city-level predictors together. The percent change in variance between empty and multivariable models was calculated to describe the total variance explained by the multivariable model. All univariable and multivariable models were adjusted for country as a fixed effect and city as a random intercept.

Data processing and analyses were conducted in R version 4.1.0(25).

## 3. Results

### 3.1. Population and urban characteristics of study area

We examined 47,187 neighborhoods in 326 cities in eight Latin American countries (Table 1). The geographic locations of observed cities are presented in Figure 1. Within study cities, urban neighborhoods in Central American cities (median [interquartile range (IQR)] NDVI, 0.59 [0.32]) and Brazil (0.57 [0.34]) were greenest while neighborhoods in Chilean cities were the least green (0.32 [0.26]). Neighborhoods in Colombia had the highest population density (12.7 [16.9] thousand residents per km^2^) while neighborhoods in Brazil were the least dense (3.9 [7.1]). At the city level, Central American cities had the greatest traffic congestion (36% [40%] longer trip duration than free-flow conditions due to congestion) while cities in Brazil were the least congested (9% [6%] longer trip duration due to congestion). Neighborhood and city population and characteristics are described overall and by country in Table 1.

**Table 1.** Population characteristics and urban form of study neighborhoods (N=47,187) within 326 Latin American cities. Results presented as number (N) or Median (interquartile range).

|  | **Total** | **Argentina** | **Brazil** | **Central America<sup>a</sup>** | **Chile** | **Colombia** | **Mexico** |
| --- | --- | --- | --- | --- | --- | --- | --- |
| **Neighborhood level** |  |  |  |  |  |  |  |
| N of neighborhoods | 47,187 | 2,044 | 4,004 | 5,563 | 744 | 3,064 | 31,768 |
| Population density<sup>b</sup> | 5.7 (9.0) | 6.5 (7.4) | 3.9 (7.1) | 6.1 (12.4) | 8.0 (7.9) | 12.7 (16.9) | 5.5 (8.2) |
| Education (% primary school) | 89.7 (12.9) | 90.8 (11.0) | 67.4 (14.9) | 78.6 (31.9) | 91.6 (8.0) | 81.5 (20.6) | 91.7 (8.8) |
| Intersection density<sup>c</sup> | 132 (127) | 87 (49) | 88 (91) | 76 (129) | 170 (133) | 111 (310) | 154 (117) |
| Greenness (NDVI) | 0.45 (0.31) | 0.41 (0.29) | 0.57 (0.34) | 0.59 (0.32) | 0.32 (0.26) | 0.51 (0.3) | 0.41 (0.3) |
| Distance from city center (km) | 8.4 (11.6) | 15.7 (23.7) | 10.3 (15.5) | 9.3 (8.9) | 4.8 (7.5) | 4.7 (6.6) | 8.2 (11.4) |
| **City level** |  |  |  |  |  |  |  |
| N of cities | 326 | 22 | 152 | 6 | 21 | 33 | 92 |
| City population (thousands) | 307 (462) | 332 (433) | 255 (402) | 1,153 (2,172) | 243 (208) | 369 (463) | 396 (655) |
| GDP per capita (USD thousands) | 14.8 (10.2) | 19.6 (11.0) | 19.3 (12.4) | 15.8 (13.3) | 17.7 (13.6) | 11.8 (4.9) | 14.0 (6.1) |
| Traffic congestion index<sup>d</sup> | 11% (11) | 10% (4) | 9% (6) | 36% (40) | 27% (12) | 32% (13) | 13% (10) |
| Population density<sup>b</sup> | 6.8 (3.4) | 5.5 (1.8) | 6.2 (2.7) | 7.9 (1.0) | 7.1 (2.3) | 15.9 (5.8) | 6.5 (2.2) |
| Education (% primary school) | 73.0 (16.5) | 79.8 (3.9) | 65.7 (8.1) | 86.7 (11.1) | 89.8 (3.8) | 83.7 (4.2) | 79.6 (8.6) |
| Intersection density<sup>c</sup> | 87.9 (33.4) | 81.2 (22.3) | 82.2 (21.1) | 60.9 (12.2) | 120.5 (19.2) | 121.6 (31.1) | 92.9 (30.8) |
| Greenness (NDVI) | 0.82 (0.10) | 0.79 (0.15) | 0.82 (0.08) | 0.88 (0.05) | 0.75 (0.51) | 0.86 (0.03) | 0.76 (0.23) |

<sup>a</sup>Central America grouping includes urban neighborhoods in Costa Rica (N=1 city), Guatemala (N=2), and Panama (N=3)
<sup>b</sup>Thousands of residents per square kilometer
<sup>c</sup>Intersections per square kilometer of built-up area
<sup>d</sup>Percent longer trip duration due to traffic congestion, as a percentage of trip time without congestion

**Figure 1.**
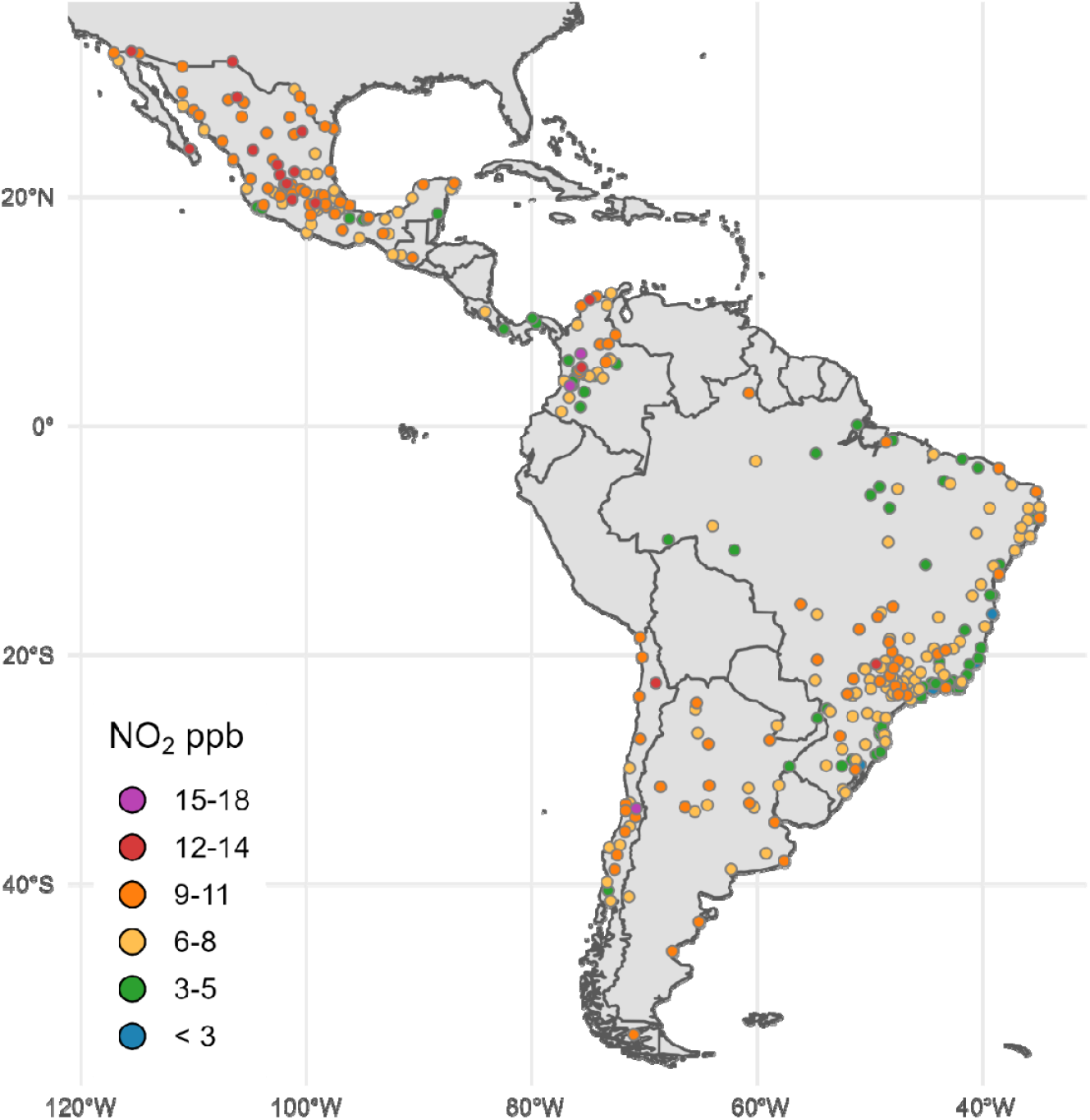
Location of study cities (N=326) and city-level population-weighted annual concentration of ambient NO_2_ in 2019. The WHO guideline for annual NO_2_ is 10 μg/m^3^ (≈ 5.3 ppb) and all neighborhoods that exceed this guideline are represented by yellow, orange, red, or purple.

### 3.2 Ambient NO_2_ neighborhood concentrations and population exposures

In Figure 1, we present the population-weighted city-level mean NO_2_ for context (as distinct from the primary analysis of neighborhood-level concentrations). City-level NO_2_ concentrations appear to have strong variation between many cities in close proximity, suggesting the importance of local drivers of ambient NO_2_. In Figure 2, we present neighborhood-level NO_2_ for two selected cities of varying sizes: the metropolitan area of Buenos Aires, Argentina (population ≈ 16 million; Panel A) and Quetzaltenango, Guatemala (population ≈ 295,000; Panel B). In both selected cities, as typical across study cities, neighborhood NO_2_ concentrations trend higher with greater proximity to the urban core. Consistent with the overall findings of Figures 1 2, our variance decomposition model showed that 9.4% of total variance in neighborhood NO2 was between countries, 30.3% of variance was between cities, and 60.3% of variance was within cities.

**Figure 2.**
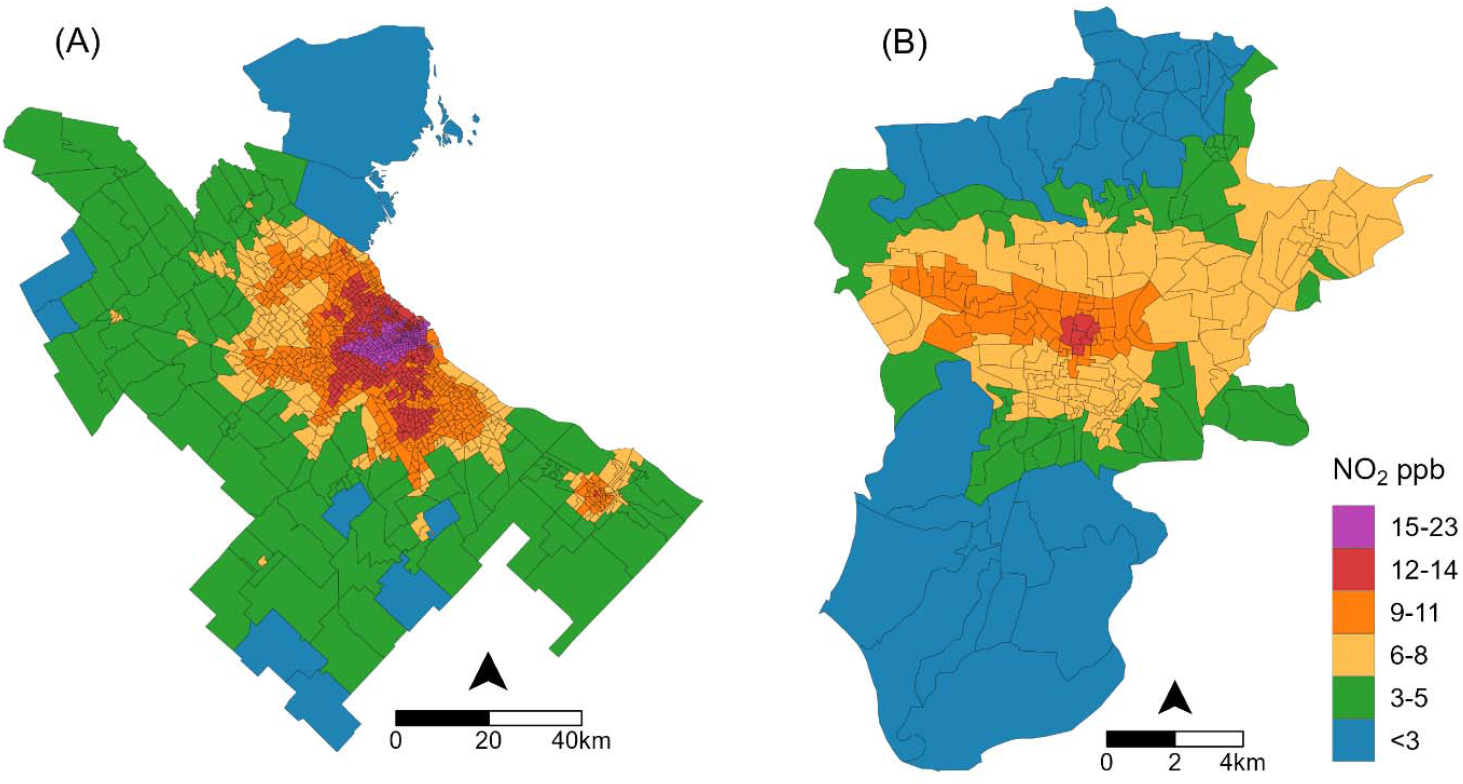
Within-city variation in neighborhood-level ambient NO_2_ in two selected cities with varying population and geographic sizes (note the panel-specific scale bars). Gray lines represent neighborhood boundaries and colors represent annual mean ambient NO_2_ in 2019. In Panel A, the metropolitan area of Buenos Aires, Argentina (population ≈ 16 million). In Panel B, Quetzaltenango, Guatemala (population ≈ 295,000). In both cities, neighborhood NO_2_ concentrations trend higher with greater proximity to the urban core. The WHO guideline for annual NO_2_ is 10 μg/m^3^ (≈ 5.3 ppb) and all neighborhoods that exceed this guideline are represented by yellow, orange, red, or purple.

The median neighborhood annual NO_2_ ppb across all countries was 10.2 ppb, nearly twice the WHO annual guideline of 5.3 ppb (Table 2). Median neighborhood NO_2_ varied between countries, ranging from 8.4 ppb in Brazil to 10.9 ppb in Argentina. We observed substantial variation in NO_2_ concentrations within countries (Table 2 and Figure 3). For example, in Colombia, the 5^th^ percentile neighborhood NO_2_ concentration was 1.4 ppb while the 95^th^ percentile concentration was 21.0 ppb. Across all countries a large proportion of neighborhoods exceeded WHO guidelines. The highest proportion of neighborhoods exceeding the guidelines was observed in Chile (95% of neighborhoods had NO_2_ concentrations above guidelines and 5% of neighborhoods had NO_2_ concentrations approximately four times greater than guidelines) and lowest in Central America (70.8% of neighborhoods exceeded guidelines).

**Table 2.** Neighborhood ambient NO_2_ concentrations and population exposures among 47,187 study neighborhoods in 326 Latin American cities.

| Country | Study Population (millions) | Study population above NO<sub>2</sub> guidelines<sup>a</sup> (millions) | Percent of study population above NO<sub>2</sub> guidelines<sup>a</sup> | Mean neighborhood NO<sub>2</sub> ppb<sup>b</sup> | 5<sup>th</sup> percentile neighborhood NO<sub>2</sub> ppb | Median neighborhood NO<sub>2</sub> ppb | 95<sup>th</sup> percentile neighborhood NO<sub>2</sub> ppb |
| --- | --- | --- | --- | --- | --- | --- | --- |
| Total | 236.0 | 199.5 | 84.6 % | 10.3 | 3.6 | 10.2 | 17.2 |
| Argentina | 23.7 | 21.9 | 91.9 % | 10.3 | 2.9 | 10.9 | 16.1 |
| Brazil | 108.4 | 84.6 | 78.0 % | 8.7 | 2.2 | 8.4 | 16.2 |
| Central America<sup>c</sup> | 6.6 | 4.7 | 70.8 % | 9.0 | 2.8 | 9.2 | 16.2 |
| Chile | 2.8 | 2.7 | 97.2 % | 11.6 | 5.9 | 10.6 | 20.8 |
| Colombia | 20.4 | 17.4 | 85.5 % | 10.3 | 1.4 | 9.4 | 21.0 |
| Mexico | 74.0 | 68.2 | 92.2 % | 10.7 | 4.9 | 10.5 | 17.1 |

<sup>a</sup> WHO annual NO<sub>2</sub> guidelines (10 µg/m<sup>3</sup> ≈ 5.3 parts per billion)
<sup>b</sup> ppb = parts per billion
<sup>c</sup> Central America grouping includes urban neighborhoods in Costa Rica (N=1 city), Guatemala (N=2), and Panama (N=3)

**Figure 3.**
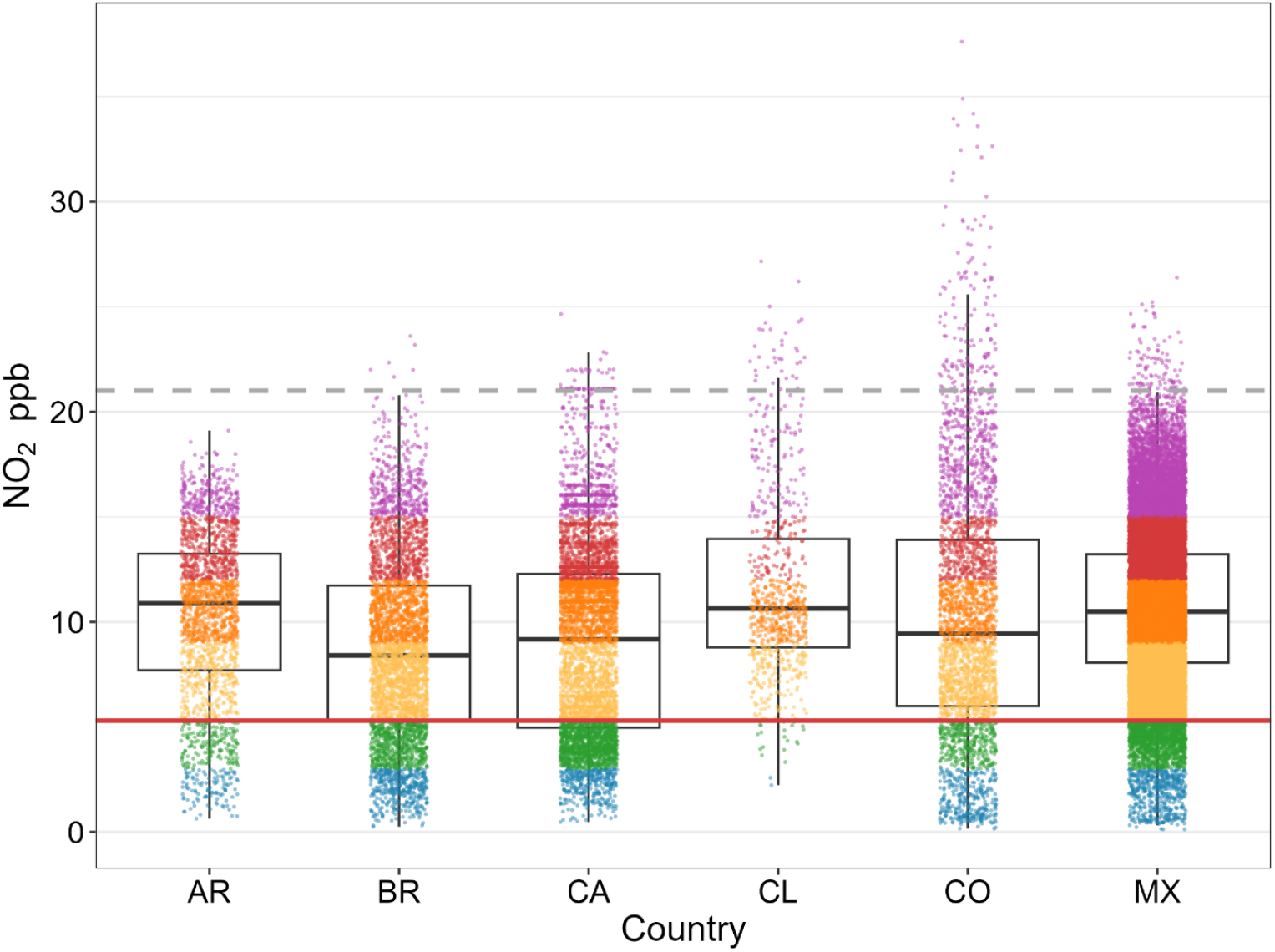
Annual ambient NO_2_ within 47,187 urban neighborhoods in Latin America. Each dot represents annual NO_2_ in one neighborhood. The red horizontal line represents the 2021 WHO guidelines for annual NO_2_ (10 μg/m^3^ ≈ 5.3 ppb). The grey dashed line represents the pre-2021 guideline for annual NO_2_ (40 μg/m^3^ ≈ 21 ppb), for reference. AR=Argentina; BR=Brazil; CA=cities in the Central American countries of Costa Rica, Guatemala, and Panama; CL=Chile; CO=Colombia; MX=Mexico.

Our study area included ≈236 million residents, ranging from 2.8 million residents in Chile to 108.4 million residents in Brazil (Table 2). Of these ≈236 million residents, nearly 200 million people (84.6% of total residents) lived with annual ambient NO_2_ concentrations above WHO guidelines. The percentage of residents living with ambient NO_2_ levels above guidelines varied from 97.2% of residents in Chilean cities to 70.8% of residents of Central American cities. All other countries had large majorities of residents living with ambient NO_2_ that exceeded guidelines (Mexico 92.2% of residents, Argentina 91.9%, Colombia 85.5%, and Brazil 78.0%).

### 3.3. Population and urban characteristics associated with NO_2_ exposure

Table 3 shows associations of neighborhood and city-level characteristics with neighborhood NO_2_ ambient concentrations. In the multilevel model adjusting for both neighborhood and city characteristics, higher neighborhood population density (0.06 higher NO_2_ ppb per unit higher population density z-score [95% confidence interval (CI) 0.04 to 0.08 ppb]) and higher educational attainment (0.64 ppb per unit higher education z-score [95% CI 0.61 to 0.67]) were associated with higher neighborhood NO_2_. Conversely, lower neighborhood NO_2_ was associated with more greenness (-2.22 ppb per unit NDVI z-score [95% CI -2.25 to -2.19]) and greater distance from the city center (-0.87 ppb per unit distance z-score [95% CI -0.90 to -0.85]). In this same model, higher city-level population density, population size, and traffic congestion were associated with higher NO_2_. Our comparison of empty and multivariable models (both adjusted for country group) indicated that 69% of the between-city variability and 79% of the between-neighborhood variability was explained by the full host of predictors.

**Table 3.**
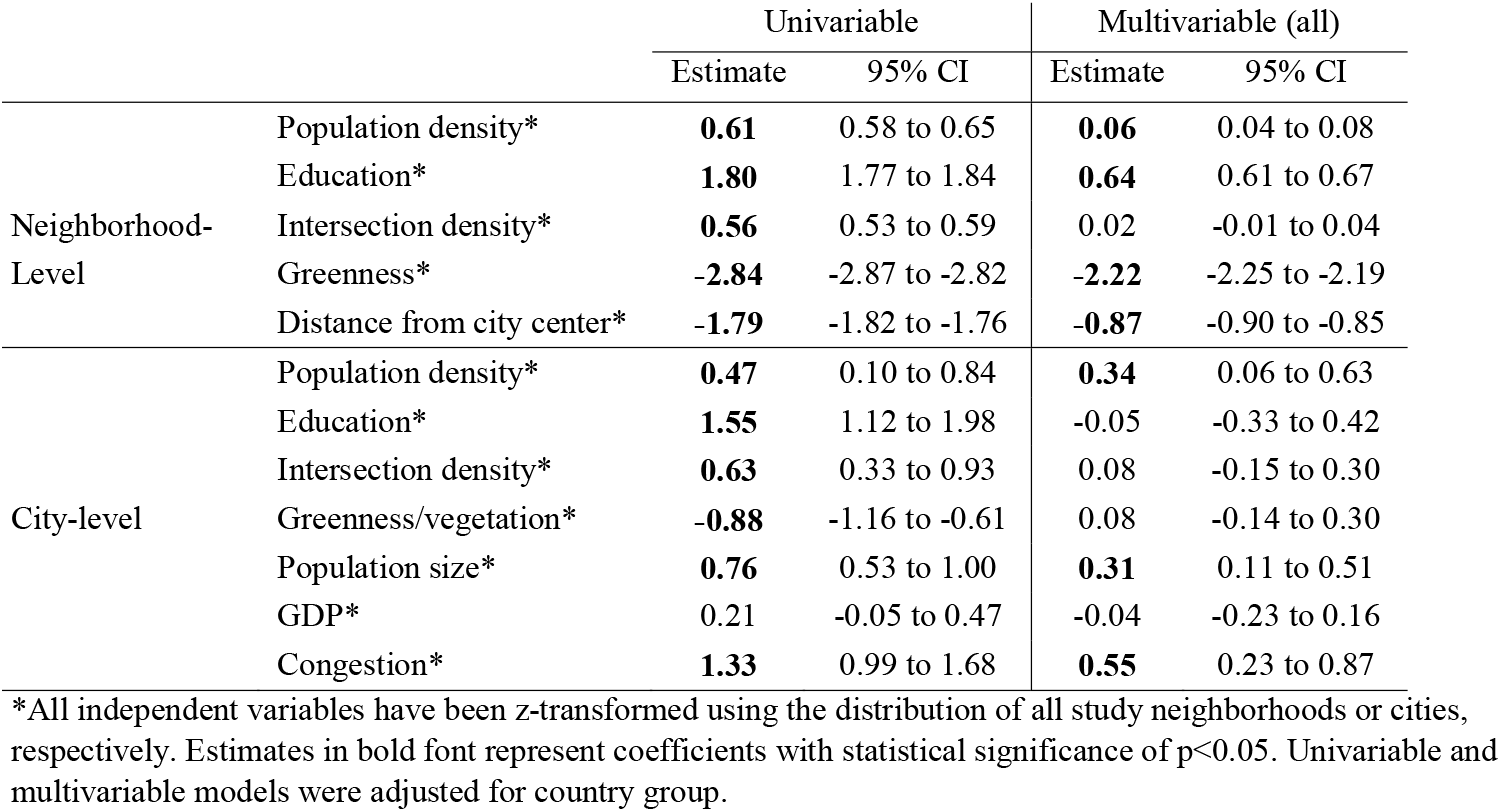
Mean differences in neighborhood ambient NO_2_ concentration (ppb) associated with a one-unit z-score increase in neighborhood- and city-level features in 47,187 urban neighborhoods in Latin America.

## 4. Discussion

We performed a highly spatially resolute descriptive analysis among ≈236 million residents of over 47,000 urban neighborhoods in 326 Latin America cities, examining 1) population exposure to ambient NO_2_ and 2) associations between neighborhood ambient NO_2_ concentrations and population characteristics and urban form. We found four key findings. First, nearly nine out of 10 residents, or around 200 million people, are exposed to ambient NO_2_ concentrations that exceed the current WHO guidelines. Second, we found that NO_2_ variability was widest within cities rather than between cities or countries. Third, larger, denser, and more congested cities had higher NO_2_. Last, within cities, we found that neighborhoods with less vegetation and closer to the city center had higher NO_2_. These findings highlight the magnitude of harmful human exposure to ambient NO_2_ in cities across Latin America, reveal important within-city differences in NO_2_ exposures, and highlight potential interventions that might reduce exposures to this harmful urban air pollutant.

We found a mean NO_2_ concentration of 10.3 ppb at the neighborhood level in 2019. We are not aware of other regional analyses of neighborhood-level NO_2_ or within-city variation of NO_2_ in Latin America, yet the concentrations we observed are similar to a city-level analysis by Anenberg and colleagues using the same NO_2_ source, that found an overall population-weighted NO_2_ concentration of 10.6 ppb across urban Latin America in the same year (14). Anenberg and colleagues estimated that urban NO_2_ across Latin America was higher than urban areas of sub-Saharan Africa (7.1 ppb) and similar to urban South Asia (10.1 ppb) and high-income countries (11.1 ppb). Given the 75% reduction in annual NO_2_ guidelines in the 2021 WHO Air Quality Guidelines(11), urban populations worldwide find themselves with long-term NO_2_ concentrations above the updated guidelines established to protect public health. Our highly spatially granular analysis of urban Latin America finds that Latin American cities are no exception, with 85% of the 236 million residents in our study area living in neighborhoods with NO_2_ concentrations above WHO guidelines.

Few studies have examined the substantial within-city variation in population exposure to ambient NO_2_ and the associations of urban and population characteristics with neighborhood differences in NO_2_ concentrations within Latin America. Understanding the drivers of between-city and within-city differences in ambient NO_2_ exposure is critical to design policies that promote health and health equity in this highly urbanized region. Our study is unique in its breadth (all cities of 100,000 residents or more in eight countries) and in our examination of how neighborhood and city level factors independently relate to urban NO_2_ exposure. We found that higher population density at both the city and neighborhood levels were independently associated with higher NO_2_. This is consistent with regional studies in the US (5) and Europe (26) and emphasizes the nature of NO_2_ pollution as a spatially-varying byproduct of local anthropogenic fossil fuel combustion. In models adjusted for city and neighborhood-level factors, we found that greenness at the neighborhood-level, but not city-level, was associated with lower neighborhood NO_2_. Overall, cities that had more vehicular traffic congestion tended to have higher NO_2_ levels. This is unsurprising given the continued dominance of fossil fuels for motor vehicles and transit, and the role of fossil fuel motorized transit in generating NO2 (1). Taken together these findings suggest that reducing city-level congestion (of fossil-fuel powered vehicles) and increasing neighborhood greenness warrant further attention as potential actionable environmental interventions to reduce population exposure to NO_2_ in urban areas.

Our findings emphasize the need for context-specific analyses of pollution exposures among populations in the Global South, where population patterns in urban areas and environmental justice concerns may be distinct from better-studied cities in high-income countries(27). Specifically, our unexpected finding of a positive association between neighborhood educational attainment and neighborhood ambient NO_2_ (i.e., neighborhoods with higher education experience higher NO_2_ concentrations). This corroborates a study of São Paulo(28), but contrasts with multicity studies of neighborhood ambient NO_2_ in Europe(29) and the United States(5). In these high-income settings, ambient NO_2_ is often higher in lower SES areas, driven largely by neighborhood urbanicity and proximity to highways. Contrasting associations between neighborhood SES and ambient air pollutants may reflect different patterns of residential segregation, specifically how segregation by SES is distributed between the urban core and urban periphery. These patterns of residential segregation by urbanicity may be evolving over time, and there is evidence in some Latin American countries that wealthier urban residents are migrating away from city centers to peripheral “private” neighborhoods outside the city center (30). However, in many Latin American cities, regardless of air pollution in their residential neighborhoods, individuals with lower SES experience substantially higher personal exposure to air pollution due to longer times spent commuting on roads with high levels of traffic-related air pollutants (31), such as NO_2_. Furthermore, individuals living in lower SES areas may experience greater health impacts of air pollution compared to those in higher SES areas due to a higher prevalence of chronic conditions and lower access to medical care(27, 32). Quantifying population exposures at the place of residence is only one step in the process of understanding the true impact of NO_2_ exposure on health disparities (27).

While we examined congestion at the city level and intersection density at the city and neighborhood levels, we were unable to look at traffic volume within the city or differences in vehicle fleet age or fuel type, which is a major driver of intra-urban variation in NO_2_ at the neighborhood level(33). Fossil-fuel powered vehicles are a major source of ambient NO_2_ and limiting their use through policies that support electric vehicles and public transit warrants critical attention as potential interventions to reduce urban NO_2_. We were also unable to include measures of indoor exposures to NO_2_ generated by indoor or household burning of fossil fuels, such as gas appliances, which is a major source of NO_2_ exposure (34, 35). In Latin America, indoor air pollution exposure from household cooking and heating varies by urbanicity and socioeconomic status (35, 36, 37) and it is likely that there are intra- and inter-urban differences in personal exposure to NO_2_ which are not captured by our measures of residential ambient NO_2_. Furthermore, while we examined neighborhood ambient NO_2_ and urban form from 2019, we used neighborhood-level population data from the most recent census for each country, with a mode census year of 2010 (Supplemental Table S1). Given larger regional trends in population growth(38) and urbanization(12), it is plausible that our estimates of population exposures based on census data are underestimates in this urbanizing region.

Our study reports on an unprecedented analysis of over 47,000 neighborhoods across this highly urbanized region of the Global South. By compiling small area census records with a new, spatially resolved estimate of ambient NO_2_, this analysis provides novel and practical evidence of within-city differences in neighborhood-level population exposure to NO_2_ in the context of recently updated WHO guidelines. This evidence can support urban policymakers and practitioners by guiding the development of policies and interventions that reduce urban NO_2_ exposure by targeting specific features of the urban environment in Latin America and beyond.

## Conclusion

Among 236 million residents of over 47,000 neighborhoods in 326 Latin American cities, nearly 9 in 10 people live in neighborhoods with ambient NO_2_ levels that exceed WHO guidelines. Neighborhoods that are denser, closer to the urban core, and have less vegetation have higher levels of NO_2_, compared to less dense neighborhoods in the urban periphery. Cities with higher vehicle congestion, population size, and population density have higher levels of ambient NO_2_. Our findings suggest that 1) increasing neighborhood-level greenness and 2) reducing city-level pollution from fossil fuel-powered vehicles by promoting active and public transit and vehicle fleet electrification have potential as actionable interventions to reduce ambient NO_2_ exposures in Latin American cities.

## Data Availability

Vital registration and population data for Brazil, Chile and Mexico were downloaded from publicly available repositories of statistical agencies in each country. Vital registration and population data for Argentina, Costa Rica, Guatemala and Panama were obtained directly from statistical agencies in each country. A link to these agency websites can be accessed via https://drexel.edu/lac/data-evidence/data-acknowledgements. Global NO2 concentrations are available at: https://doi.org/10.6084/m9.figshare.12968114 as described here: https://doi.org/10.1016/S2542-5196(21)00255-2. Please contact the corresponding author about access to neighborhood level urban features.

## Acknowledgments and funding sources

This study was financially supported by the Wellcome Trust [205177/Z/16/Z]. The authors acknowledge the contribution of all SALURBAL project team members. For more information on SALURBAL and to see a full list of investigators, see https://drexel.edu/lac/salurbal/team/. SALURBAL acknowledges the contributions of many different agencies in generating, processing, facilitating access to data or assisting with other aspects of the project. Please visit https://drexel.edu/lac/data-evidence for a complete list of data sources. JLK was supported by the Drexel FIRST (Faculty Institutional Recruitment for Sustainable Transformation) Program, National Institutes of Health grant number U54CA267735-02, and the Cotswold Foundation Postdoctoral Fellowship Grant# 230356. UB was also supported by Office of the Director of the National Institutes of Health under award number DP5OD026429. The funding sources had no role in the analysis, writing, or decision to submit the manuscript.

## Author contributions

JLK, UB, and AVDR conceptualized the analysis. AVDR acquired funding. AVDR and UB provided supervision. JLK curated data, completed the formal analysis, and created the original draft of the manuscript. All authors had access to underlying data and contributed to methodology and manuscript review and editing.

