## Supplementary Material for "Ambient nitrogen dioxide in 47,187 neighborhoods across 326 cities in eight Latin American countries: population exposures and associations with urban features"

Supplementary Information

**Supplemental Table S1**: Neighborhood administrative units and years of most recent available census.

| **Country** | **Administrative unit (neighborhood)** | **Census/population year** |
| --- | --- | --- |
| Argentina | Fraccion Censal | 2010 |
| Brazil | Áreas de Ponderação | 2010 |
| Chile | Zona Censal | 2017 |
| Colombia | Sector Urbano, Clase = 1 | 2018 |
| Costa Rica | Distrito | 2011 |
| Guatemala | Sector Censal | 2002 |
| Mexico | Área Geoestadistica Básica | 2010 |
| Panama | Barrio | 2010 |
